# Respiratory symptoms after coalmine fire and pandemic: a longitudinal analysis of the Hazelwood Health Study adult cohort

**DOI:** 10.1101/2023.08.23.23294510

**Authors:** Tyler J. Lane, Matthew Carroll, Brigitte M. Borg, Tracy A. McCaffrey, Catherine L. Smith, Caroline X. Gao, David Brown, Amanda Johnson, David Poland, Shantelle Allgood, Jillian Ikin, Michael J. Abramson

## Abstract

**Objectives:** Extreme but discrete fine particle <2.5μm (PM_2.5_) exposure is associated with higher prevalence of respiratory symptoms. It was unknown whether these effects abate, persist, or worsen over time, nor whether they are exacerbated by COVID-19.

**Methods:** We analysed longitudinal survey data from a cohort residing near a 2014 coalmine fire in regional Australia. A 2016/2017 survey included 4,056 participants, of whom 612 were followed-up in 2022. Items included questions on respiratory symptoms, history of COVID-19, and time-location diaries that were combined with geospatial and temporal models of fire-related PM_2.5_. Associations were examined using logistic and mixed-effects logistic regressions.

**Results:** PM_2.5_ exposure predicted higher prevalence of chronic cough and current wheeze 2-3 years post-fire. At the 2022 follow-up, PM_2.5_ exposure was associated with worsening prevalence of chronic cough and possibly current wheeze. While there were no detectable interaction effects between PM_2.5_ and COVID-19, PM_2.5_ exposure was associated with additional respiratory symptoms among participants who reported a history of COVID-19.

**Conclusion:** Short-term but extreme PM_2.5_ may increase the long-term prevalence of chronic cough, while COVID-19 may exacerbate the effect on additional respiratory symptoms.

## 1 Introduction

Exposure to fine particulate matter <2.5 μm (PM_2.5_) worsens respiratory health (1,2). When the source is an extreme combustion event like wildfire, PM_2.5_ is likely even more harmful (3). In early 2014, a coalmine fire near the Hazelwood power station in regional Victoria, Australia, shrouded the adjacent town of Morwell in smoke and ash for six weeks. Daily mean PM_2.5_ reached 1022μg/m^3^ in residential areas closest to the mine (4), nearly seven times the Environment Protection Authority Victoria’s threshold of 150μg/m^3^ for “extremely poor” air quality (5). Those residing in Morwell reported numerous symptoms during the coalmine fire including respiratory problems like shortness of breath, chest tightness, and cough (6).

The Hazelwood Health Study was established to investigate how smoke from the coalmine fire affected long-term health. In the years since, it has found numerous associations between PM_2.5_ exposure and poorer respiratory health. PM_2.5_ during the fire was associated with increased use of respiratory medical services including GP and specialist visits (7), hospital admissions and emergency presentations (8), and dispensing of inhaled medicines (9). There is evidence that effects have persisted for at least a few years, including higher prevalence of respiratory symptoms measured between 2-4 years after the fire (10,11), accelerated lung ageing, features of chronic obstructive pulmonary disease (COPD) at 4 years (11–13), and increased respiratory emergency department presentations in the five years post-fire (14). Recent findings suggest some recovery in lung function 7.5 years after the fire (15), though the long-term effects remain unclear.

Late 2019 saw the emergence of another broad threat to respiratory health, the COVID-19 pandemic. Both particulate matter and COVID-19 can increase inflammatory immune responses in the lungs (2,16). PM_2.5_ exposures may even increase the likelihood that SARS-CoV-2 infection leads to a cytokine storm (17), or sustained inflammation that can damage the respiratory system (16). The combination of ambient PM_2.5_ exposure and COVID-19 has been associated with more severe disease (18). However, it was unknown whether discrete but extreme PM_2.5_ exposure from smoke events have similar exacerbating effects when they have occurred several years in the past (19).

In this analysis, we addressed the following research questions:

1. Does coalmine fire-related PM_2.5_ affect long-term respiratory symptom trajectory?
2. Does COVID-19 infection worsen the long-term effects of coalmine fire-related PM_2.5_ exposure on respiratory symptoms?

## 2 Methods

This study was pre-registered on the Open Science Framework on 19 July 2022, the month before the follow-up survey commenced, and updated during the survey with a more specific analysis plan on 22 November 2022 (20).

### 2.1 Participants

This analysis uses data from two surveys of the Hazelwood Health Study’s Adult Cohort. The cohort was established in the initial survey, administered from May 2016 to February 2017. Using electoral rolls, we identified individuals who had been residing in Morwell, a town adjacent to the coalmine and whose residents experienced the most smoke exposure, and the residents from the nearby unexposed site of Sale, during the coalmine fire (see Figure 1) and invited them to participate. This resulted in a cohort of 4,056 (21). From August to December 2022, we sent emails and mobile phone invites to cohort members if they 1) had provided email/mobile numbers, 2) were not part of a separate follow-up survey on mental health outcomes, 3) consented to further contact to participate in a follow-up study, and 4) not known to be dead. Data were collected using REDCap (22,23).

**Figure 1.**
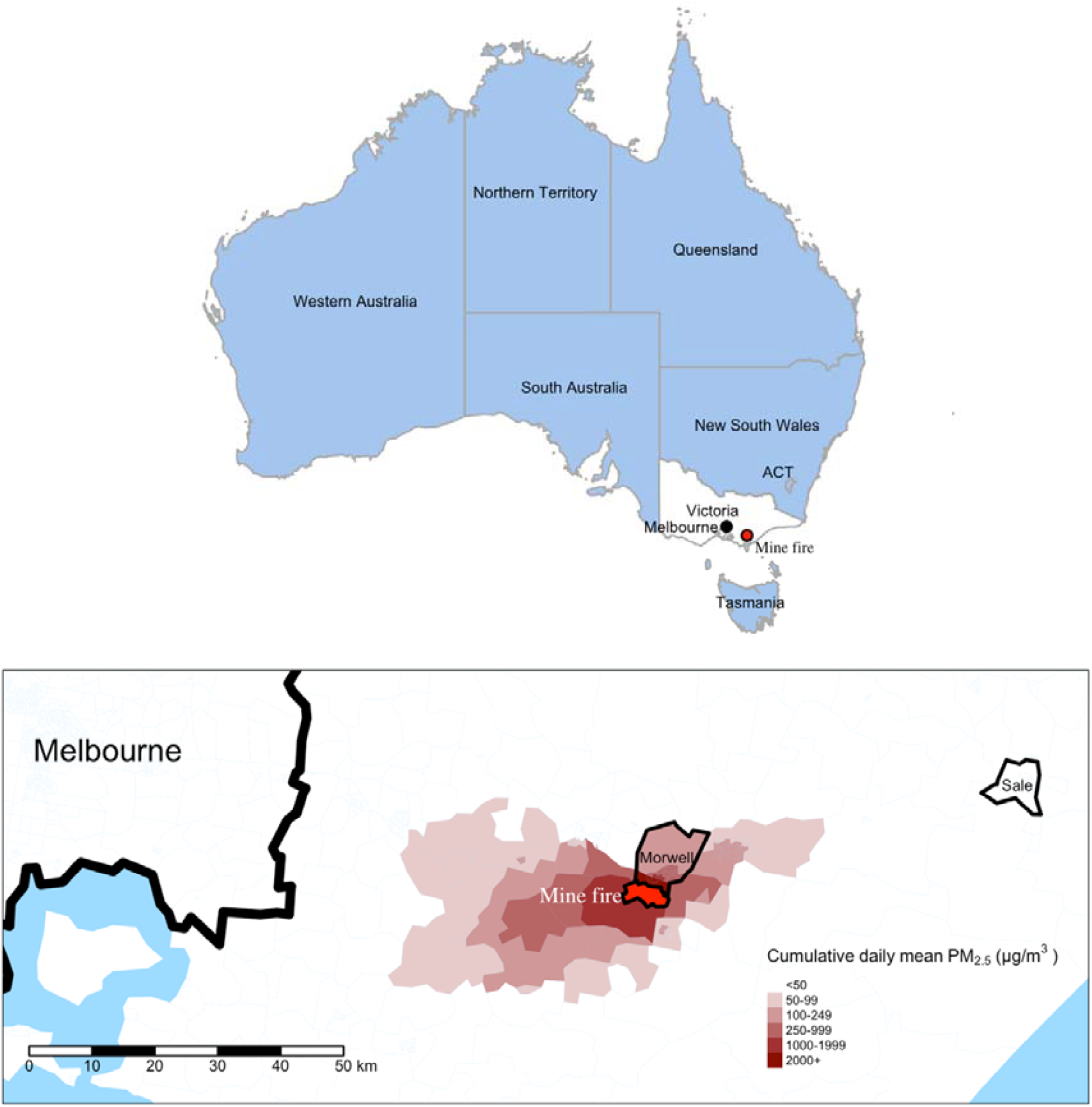
Maps of Australia illustrating location of the coalmine and distributions of fire related-PM_2.5_ (February to March 2014) in the surrounding areas, highlighting exposure site Morwell and control site Sale

#### 2.1.1 Exposures

Participant’s daily mean PM_2.5_ exposure during the coalmine fire was determined by blending self-reported time-location diaries, which placed individuals throughout the mine fire period at 12-hour intervals, with modelled estimates of hourly coalmine fire-related PM_2.5_ concentrations (21). Estimated PM_2.5_ concentrations were generated using chemical transport models, incorporating air monitoring data, coal combustion, and weather conditions. Spatial models achieved a resolution of 100m^2^ in the area surrounding the mine fire (surrounding 10.1km^2^ area) with precision decreasing along with distance from the fire. For more detail, refer to Luhar et al. 2020 (4), particularly Figure 8. Cumulative daily mean fire-related PM_2.5_ distributions are illustrated in Figure 1, showing the locations of the coalmine, exposure site Morwell, and control site Sale.

History of COVID-19 was self-reported using a standardised questionnaire (24), available in the supplementary materials.

#### 2.1.2 Outcomes

Respiratory symptom questions were derived from a modified version of the European Community Respiratory Health Survey (ECRHS) Short Screening Questionnaire and included: current wheeze, chest tightness, nocturnal shortness of breath, resting shortness of breath, current nasal symptoms, chronic cough, and chronic phlegm (25). Using chronic cough and chronic phlegm responses, we created two additional outcomes: chronic wet cough (chronic cough with phlegm) and chronic dry cough (chronic cough without phlegm). Items are available in supplementary materials.

#### 2.1.3 Confounders

We adjusted for potential confounders of the effect of mine fire-related PM_2.5_ exposure and COVID-19 on the development of respiratory symptoms. Demographic factors included age at the initial (2016/17) survey, transformed with a natural spline to account for non-linear effects, and sex. Tobacco use was measured with smoking status (current, former, never) and cigarette pack-years (the number of packs of 20 cigarettes smoked per day multiplied years smoked, which was square root-transformed to account for extreme right skew. We adjusted for socioeconomic factors using the Index of Relative Socioeconomic Advantage and Disadvantage (IRSAD) score for residential area at Statistical Area Level 2 based on 2016 census data (26) and individual educational attainment. Diagnosis of asthma or COPD before the coalmine fire was treated as a confounder. For analyses of the longitudinal effects of PM_2.5_ on respiratory symptoms, COVID-19 was included as a confounder. Being fully vaccinated against COVID-19 (4+ vaccinations) was included as a confounder in all analyses.

### 2.2 Statistical analyses

Descriptive statistics were used to characterise the sample by survey round and area of residence during the coalmine fire. Continuous variables are presented as medians and interquartile ranges and categorical/dichotomous variables as counts and percentages.

To estimate the longitudinal effects of PM_2.5_ on respiratory symptoms, we conducted crude and adjusted mixed-effects logistic regression models for each outcome, using interaction terms between PM_2.5_ and survey round and random intercepts for participants. This analysis included all cohort members.

To estimate whether COVID-19 moderated the effect of PM_2.5_ on respiratory symptoms, we conducted crude and adjusted logistic regressions for each outcome, using interaction terms between PM_2.5_ and COVID-19. This analysis only included cohort members who participated in the follow-up survey. Each model adjusted for the presence of the specific respiratory symptom at the baseline survey (e.g., the model assessing whether COVID-19 exacerbated effects of PM_2.5_ exposure on prevalence of chronic cough at the 2022 follow-up survey adjusted for presence of chronic cough at the 2016/2017 baseline survey).

Residence during the mine fire (Morwell or Sale) was added to final models as this may have captured unmeasured confounding due to lifestyle, socioeconomic status, and access to health services. Sensitivity analyses on the moderating effect of COVID-19 added inverse probability weights (balanced) based on differences in confounders between responders and non-responders, using initial survey data.

To account for missing data, we used multiple imputation by chained equations and pooled results according to Rubin’s rules (27). While survey data are confidential and cannot be shared, we have archived cleaning and analytical code on a public repository (28). Data were analysed in R (29) using RStudio (30). More detail about statistical packages is available in the supplementary materials.

### 2.3 Ethics

This study received approval from the Monash University Human Research Ethics Committee (MUHREC) as part of the Hazelwood Adult Survey & Health Record Linkage Study (Project ID: 25680; previously CF15/872 – 2015000389 and 6066). Cohort members gave informed consent for each survey in which they participated.

## 3 Results

### 3.1 Descriptives

From the 4,056 members of the original cohort, 2,458 (61%) were invited to participate in the follow-up survey, of whom 612 (25%) participated. National Death Index data (31), which were linked to cohort data after the survey was completed, indicated 143 invited cohort members (7.7%) died prior to the close of the survey window (December 2022). Of the remaining 1703 non-participants, 1359 (80%) did not respond to invites, 224 (13%) refused, 68 (4.0%) gave consent but did not complete the survey, and 52 (3.1%) were not contactable. Descriptive statistics by survey round and residence can be found in Table S1 in the supplementary materials. Among Morwell respondents, the prevalence of all respiratory symptoms increased significantly between surveys, as did current wheeze, chest tightness, resting shortness of breath, and nasal allergy among Sale respondents (see Figure S1 in the supplementary materials).

### 3.2 Longitudinal effects of coalmine fire-related PM_2.5_ on respiratory symptoms

At the initial survey (2-3 years post-fire), crude and adjusted analyses found the prevalence of chronic cough was significantly higher in association with mine fire-related PM_2.5_ exposure. Prevalence of current wheeze and chest tightness were also elevated in crude and adjusted models, though both attenuated in magnitude and significance with adjustment for study site (Morwell or Sale).

Between the initial 2016/17 survey and 2022 follow-up (8.5-9 years post-fire), the prevalence of chronic cough increased in conjunction with PM_2.5_ exposure. Crude models also found current wheeze increased in association with PM_2.5_ exposure between the initial and follow-up survey. While point estimates and precision were similar in adjusted models, there was a slight attenuation to non-significance. The long-term effects of PM_2.5_ – from the 2014 coalmine fire to 2022 follow-up survey – were increased prevalence of both chronic cough and current wheeze. While effects among chronic cough subtypes were mostly non-significant, the direction and magnitude of associations indicated the increase in overall chronic cough over time was mostly attributable to wet cough. Model results are illustrated in Figure 2 and summarised in Supplementary Table 1.

**Figure 2.**
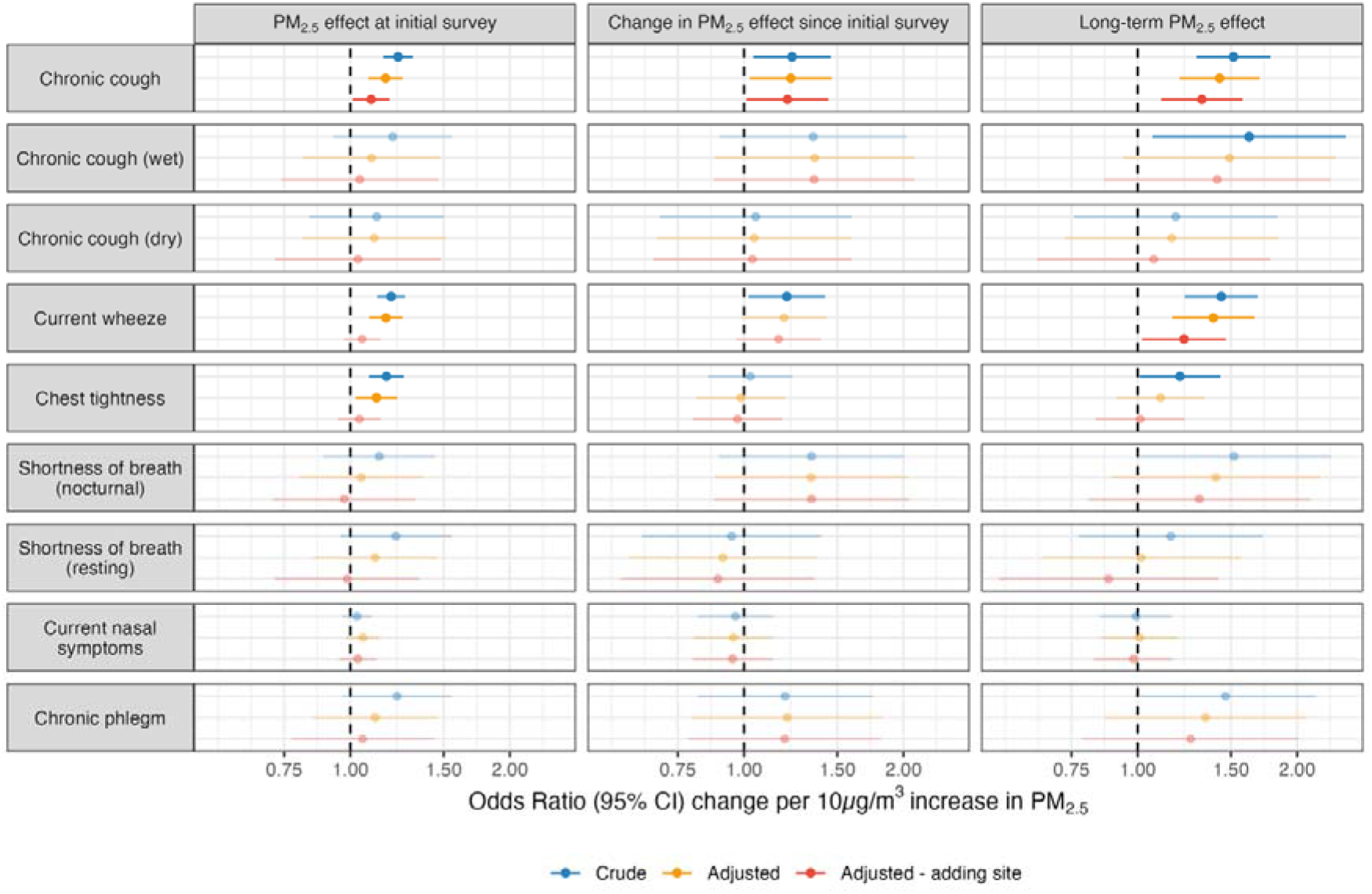
Effects of 10μg/m^3^ increase in daily mean coalmine fire-related PM_2.5_ on odds of respiratory symptoms at the initial 2016/17 survey (2-3 years post-fire), change between the two survey rounds (interaction term between fire-related PM_2.5_ and survey round), and the cumulative long-term effect at the 2022 follow-up (8.5-9 years post-fire, estimated using linear combination of PM_2.5_ and interaction term); faded points and intervals indicate non-significant effects

To investigate whether PM_2.5_ effects on wheeze were the result of worsening asthma control, we stratified analyses based on self-reported asthma diagnoses at the initial 2016/17 survey. Perhaps surprisingly, effects were isolated to non-asthmatics. There was also a total reduction in chest tightness among asthmatics. These results are presented in Figure S2 in the supplementary materials.

### 3.3 Interaction effects of PM_2.5_ and COVID-19 on respiratory symptoms

Model results from these analyses are summarised in Figure 3 and Table S3 in the supplementary materials. While interaction effects between PM_2.5_ exposure and COVID-19 were non-significant, point estimates were positive for chronic wet cough (OR range across crude, adjusted, and adjusted – adding site models: 1.22-1.31) current wheeze (OR range: 1.14-1.18), chest tightness (OR range: 1.25-1.32), nocturnal shortness of breath (OR range: 1.27-1.32), and chronic phlegm (OR range: 1.23-1.32), suggesting some exacerbation of PM_2.5_ effects by COVID-19. Conversely, interactions trended negative with chronic dry cough (OR range: 0.76-0.79) and resting shortness of breath (OR range: 0.80-0.82).

**Figure 3.**
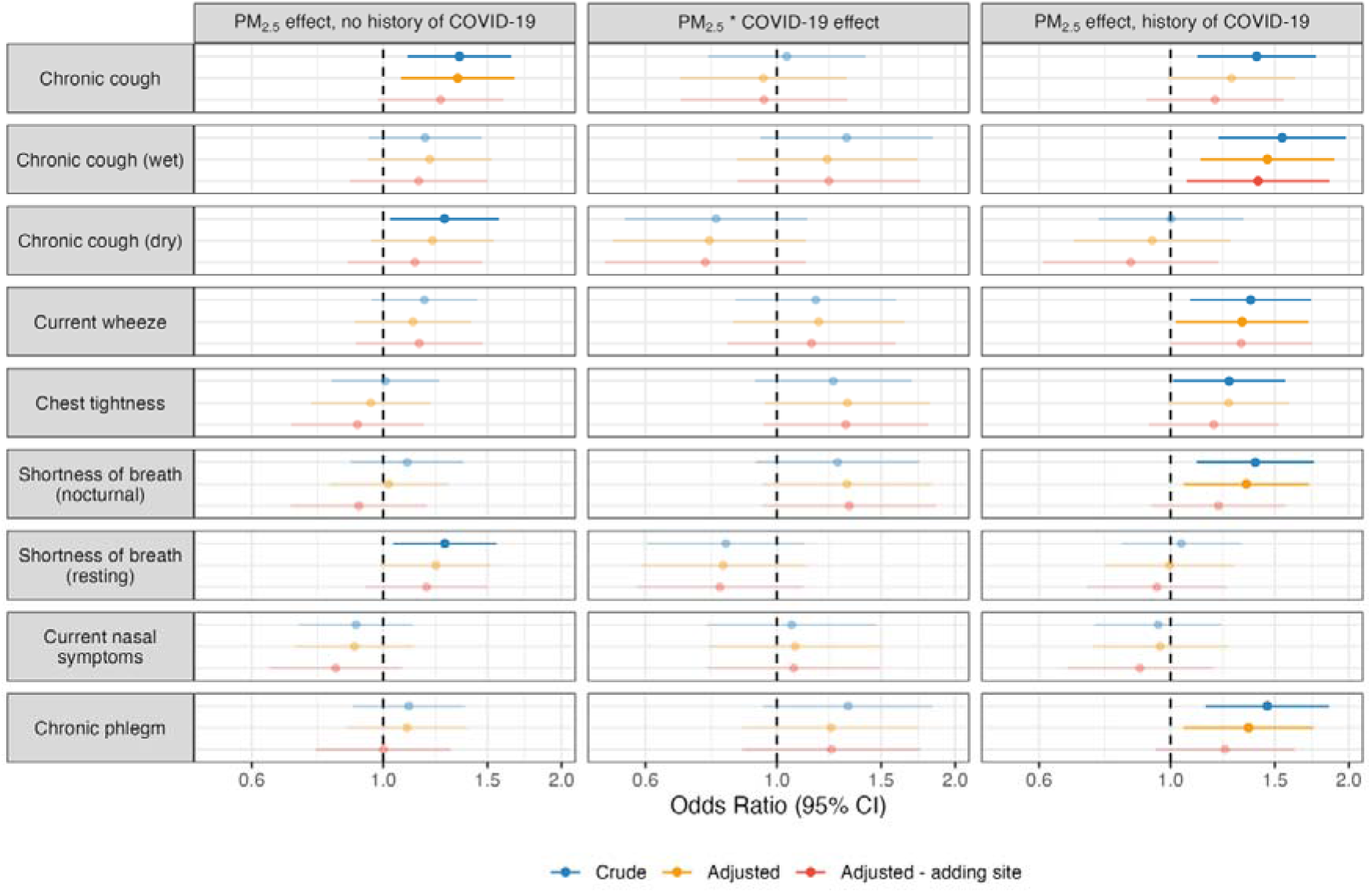
Effects of 10μg/m^3^ increases in coalmine fire-related PM_2.5_ and COVID-19 on prevalence of respiratory symptoms and whether there was a moderating effect, 8.5-9 years after the fire; faded points and intervals indicate non-significant effects

Notably, associations between PM_2.5_ and respiratory symptoms varied based on history of COVID-19. Among those with no history of COVID-19, PM_2.5_ exposure was only associated with chronic cough and possibly dry chronic cough, and resting shortness of breath. However, among those with a history of COVID-19, PM_2.5_ exposure was associated with *additional* respiratory symptoms: chronic wet cough, current wheeze, nocturnal shortness of breath, chronic phlegm, and possibly chest tightness. It is worth noting that while many of these effects attenuated to non-significance with the adjustment for study site, point estimates remained positive; only chronic wet cough was robust to adjustment for study site.

## 4 Discussion

Our first research question concerned the long-term effects of coalmine fire-related PM_2.5_ on respiratory symptoms. We found that PM_2.5_ exposure from the Hazelwood coalmine fire was associated with an increased prevalence of chronic cough, along with a possible increase in the prevalence of current wheeze. Not only was the prevalence of these conditions still elevated 8.5-9 years after the fire, they also appeared to increase over time. Chronic cough is associated with deteriorations in physical, mental and social health (32), and treatments are limited and complex (33). Therefore, these findings are a concerning development for our cohort and others around the world who have been exposed to extreme smoke events.

One possible mechanism is cough hypersensitivity, which lowers the threshold for a physiological cough response (34,35) and has previously been associated with environmental triggers such as PM_2.5_ (34). Such an effect has been demonstrated after other discrete but extreme air pollutant exposures like the 9/11 terrorist attacks in New York and earthquake rescue operations (34). The precise mechanism is unclear, though there is evidence for neuropathology such as upregulation of sensory neuroreceptors (e.g., P2X3, TRPA1, and TRPV1) in response to air pollutants (34,36) including PM_2.5_ (37). It is unclear why the effects worsened over time. The 2019/2020 Black Summer bushfires may have exacerbated the coalmine fire’s effects on the prevalence of chronic cough.

Our second research question investigated whether COVID-19 exacerbated the effects of coalmine fire-related PM_2.5_. We found some evidence of this. While interaction terms between PM_2.5_ and COVID-19 were non-significant, point estimates among several symptoms were positive and robust to adjustment, and some PM_2.5_-respiratory symptom associations (chronic wet cough, current wheeze, nocturnal shortness of breath, chronic phlegm, and possibly chest tightness) were only detectable among participants with a history of COVID-19. Exacerbation of PM_2.5_ effects by COVID-19 may also explain why the prevalence of current wheeze worsened in association with PM_2.5_. Notably, in sensitivity analyses we found that the increase in current wheeze was isolated to non-asthmatics, and there was a *reduction* in chest tightness among asthmatics. One explanation is diagnosed asthmatics have access to inhaled medications that enable them to control respiratory symptoms like wheeze, while non-asthmatics do not. To our knowledge, this is the first study to provide evidence that the effects of historical exposure to extreme but discrete smoke from a coalmine fire on respiratory symptom prevalence may be exacerbated by subsequent SARS-CoV-2 infection. However, given the tenuous nature of these associations, we highlight them with due caution.

Interestingly, fire-related PM_2.5_ exposure was associated with higher prevalence of chronic cough regardless of COVID-19 history (while only significant in the crude model among those with a history of COVID-19, point estimates only attenuated slightly with adjustment for potential confounders). Though there was limited statistical power for analysis of wet and dry chronic cough subtypes, the patterns of association suggested that contracting COVID-19 increased the likelihood that chronic cough, which may have developed due to smoke exposure, was later accompanied by phlegm, as opposed to developing a new wet cough. These association patterns included: 1) PM_2.5_*COVID-19 interaction effects trending positive for the effect on wet cough and negative for dry cough; 2) among those with a history of COVID-19, a clear positive effect of PM_2.5_ on wet cough and null effect on dry cough, with no such effect observed among those without a history of COVID-19; and 3) a null interaction between PM_2.5_*COVID-19 on overall chronic cough. It remains unclear why this happened.

### 4.1 Strengths and limitations

Among this study’s strengths are the longitudinal design, use of time-location diaries and modelled air pollution data to estimate individual-level PM_2.5_ exposure, accounting for events occurring between survey rounds like the COVID-19 pandemic, adjustment for important confounders such as socioeconomic factors and smoking, and consistent use of standardised validated measures for respiratory symptom outcomes.

The participation rate and sample size in the follow-up survey were modest. We lacked participant data from before the coalmine fire. Time-location diaries used to determine where participants were during the mine fire were likely subject to recall bias, especially since they were administered 2-3 years after the event. PM_2.5_ exposure was based on modelled estimates, which could deviate from actual concentrations (4), and do not account for individual variations due to wearing masks or time spent outdoors. The uniqueness of this cohort, particularly their older age, regional location, and residence within a developed country that was highly vaccinated before COVID-19 became widespread, may not generalise findings to other settings.

## 5 Conclusions

Our findings suggest extreme but short-term PM_2.5_ exposure from a coalmine fire has negative long-term effects on respiratory health, most noticeably chronic cough, possibly due to inducement of cough hypersensitivity. This is concerning because the effect appeared to worsen over time, and chronic cough is associated with a deterioration in health. We also found limited evidence that COVID-19 exacerbated effects of coalmine fire-related PM_2.5_.

There are serious implications for the millions of Australians affected by the 2019/2020 Black Summer bushfires, the hundreds of millions affected by 2020 and 2023 wildfires across North America and elsewhere. As climate change increases the frequency, intensity, duration, and spread of fires, smoke exposure will grow as a global public health problem, and along with habitat destruction and the wild animal trade, may be exacerbated by the spread of new zoonotic diseases.

## Data Availability

All data produced in the present study are confidential survey responses and therefore cannot be shared. However, we have provided a link to a public repository on which we have archived our cleaning and analytical code.

https://doi.org/10.26180/22596994.v4

