## supplementary materials for "Respiratory symptoms after coalmine fire and pandemic: a longitudinal analysis of the Hazelwood Health Study adult cohort"

### 1 Descriptives table

Table S1. Sample descriptive statistics, divided by residence during the mine fire (Morwell or Sale) and survey round (2016-2017 original or 2022 follow-up); count variables are presented as *n* (%) and continuous as median (IQR)

|  | **2016-2017 original survey** | | **2022 follow-up survey** | |
| --- | --- | --- | --- | --- |
|  | **Morwell**  **(*n* = 3037)** | **Sale**  **(*n* = 960)** | **Morwell**  **(*n* = 408)** | **Sale**  **(*n* = 204)** |
| Mean 24-hour PM_2.5_ exposure µg/m^3^ | 11 [IQR: 7, 19] | 0 [IQR: 0, 0] | 11 [IQR: 8, 19] | 0 [IQR: 0, 0] |
| Ever had COVID-19  *Missing* | - | - | 168 (41%)  *3* | 86 (42%)  *0* |
| Full COVID-19 vaccination (4+)  *Missing* | - | - | 238 (59%)  *3* | 104 (51%)  *1* |
| **Demographics/confounders** |  |  |  |  |
| Age at survey  *Missing* | 59 [IQR: 46, 70]  *5* | 59 [IQR: 45, 70]  *0* | 63 [IQR: 54, 71]  *1* | 64 [IQR: 55, 73]  *0* |
| Female | 1705 (55%) | 550 (57%) | 236 (58%) | 126 (62%) |
| Educational attainment  Secondary up to year 10  Secondary, years 11-12  Certificate/diploma/tertiary  *Missing* | 1006 (33%)  668 (22%)  1381 (45%)  *41* | 241 (25%)  162 (17%)  546 (58%)  *11* | 98 (24%)  80 (20%)  228 (56%)  *2* | 29 (14%)  41 (20%)  133 (66%)  *1* |
| Smoker status  Non-smoker  Previous smoker  Current smoker  *Missing* | 1495 (49%)  1052 (34%)  516 (17%)  *33* | 498 (52%)  325 (34%)  126 (13%)  *11* | 190 (47%)  167 (41%)  47 (12%)  *4* | 107 (53%)  84 (42%)  11 (5.4%)  *2* |
| Cigarette pack-years (excl. non-smokers) | 14 [IQR: 5, 30] | 15 [IQR: 5, 26] | 12 [IQR: 4, 24] | 20 [IQR: 5, 30] |
| Socioeconomic score (IRSAD)*  *Missing* | 833 [IQR: 782, 889]  *75* | 898 [IQR: 844, 952]  *0* | 838 [IQR: 782, 909]  *14* | 930 [IQR: 844,952]  *0* |
| Asthma (pre-2014)  *Missing* | 715 (23%)  *17* | 205 (21%)  *5* | 116 (29%)  *1* | 44 (22%)  *0* |
| COPD (pre-2014)  *Missing* | 148 (4.8%)  *10* | 27 (2.8%)  *4* | 11 (2.7%)  *2* | 2 (1.0%)  *0* |
| **Outcomes** |  |  |  |  |
| Chronic cough  *Missing* | 989 (32%)  *21* | 189 (20%)  *6* | 170 (42%)  *4* | 44 (22%)  *0* |
| Chronic cough (wet)  *Missing* | 600 (20%)  *28* | 107 (11%)  *6* | 92 (23%)  *6* | 24 (12%)  *0* |
| Chronic cough (dry)  *Missing* | 385 (13%)  *28* | 82 (8.6%)  *6* | 77 (19%)  *6* | 29 (9.8%)  *0* |
| Current wheeze  *Missing* | 1317 (43%)  *16* | 254 (27%)  *5* | 207 (51%)  *3* | 67 (33%)  *0* |
| Chest tightness  *Missing* | 792 (26%)  *16* | 154 (16%)  *3* | 132 (33%)  *3* | 42 (21%)  *0* |
| Shortness of breath (nocturnal)  *Missing* | 635 (21%)  *15* | 120 (13%)  *3* | 108 (27%)  *3* | 25 (12%)  *1* |
| Shortness of breath (resting)  *Missing* | 611 (20%)  *27* | 90 (9.4%)  *3* | 112 (28%)  *1* | 32 (16%)  *0* |
| Current nasal symptoms  *Missing* | 1175 (38%)  *18* | 347 (36%)  *5* | 200 (49%)  *3* | 89 (44%)  *0* |
| Chronic phlegm  *Missing* | 785 (26%)  *21* | 150 (16%)  *3* | 115 (28%)  *2* | 31 (15%)  *0* |

*Index of Relative Socioeconomic Advantage and Disadvantage (IRSAD) score, from the Socio-Economic Indexes for Areas (SEIFA)^1^

#### 1.1 Changes in respiratory symptoms between surveys

Using a crude mixed-effects logistic regression, we compared the prevalence of respiratory symptoms between the initial and follow-up surveys between study sites. While the prevalence of each symptom increased in Morwell, in Sale there were increases in four of the nine symptoms: current wheeze, chest tightness, resting shortness of breath, and nasal allergy. These effects are summarised in Figure S1.

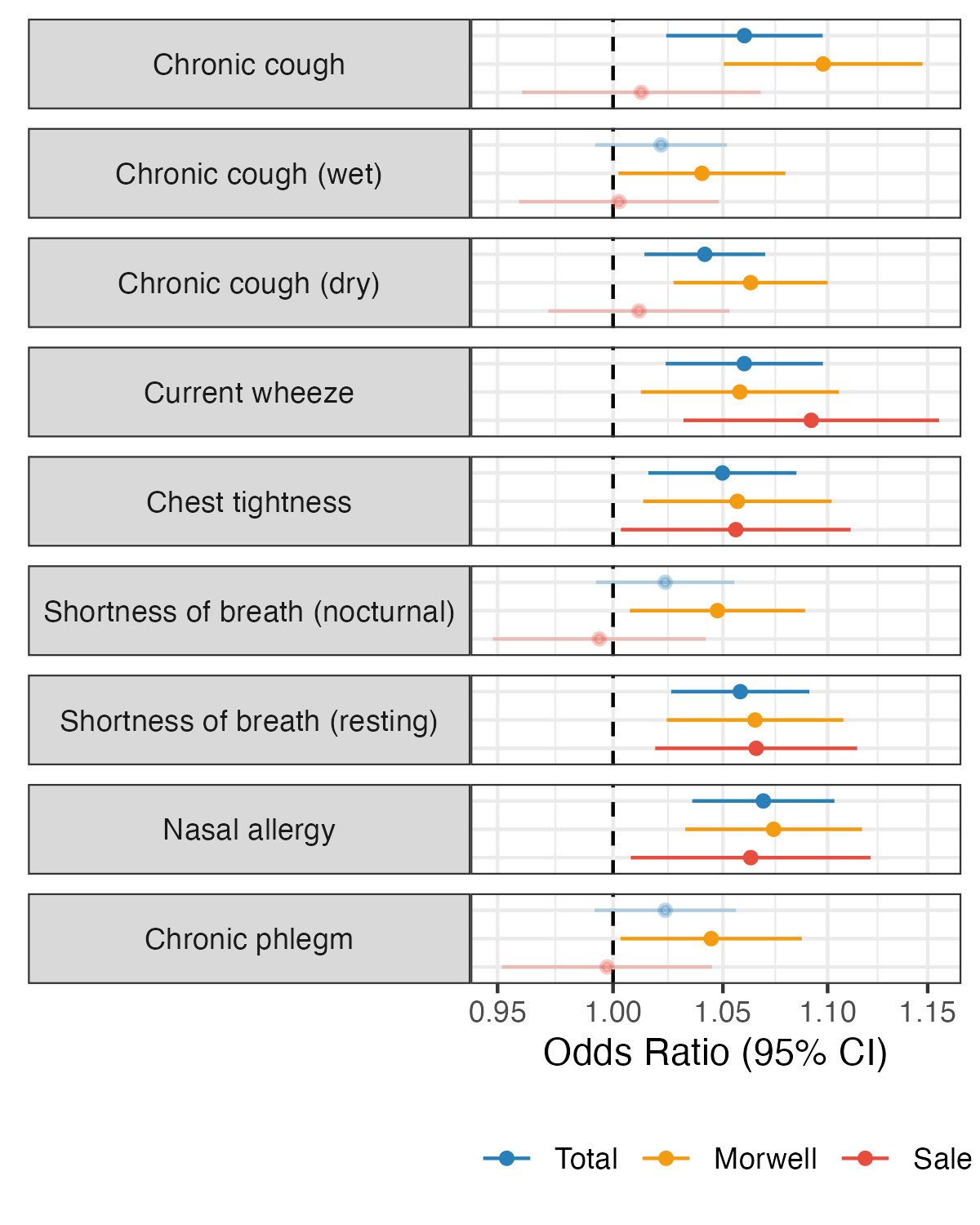

Figure S1. Changes in the prevalence of respiratory symptoms between the initial 2016/17 survey (3-4 years post-fire) and the 2022 follow-up (8.5-9 years post-fire)

### 3 Model output tables

Table S2. Effects of 10µg/m^3^ increase in mean coalmine fire-related PM_2.5_ on odds of respiratory symptoms at the initial 2016/17 survey and changes to the 2022 follow-up; significant associations at *p* ≤ 0.05 in bold

| **Respiratory symptom** | **Model type** | **PM_2.5_ effect at initial survey** | **Change in PM_2.5_ effect since initial survey** | **Long-term PM_2.5_ effect** |
| --- | --- | --- | --- | --- |
| **Chronic cough** | Crude | **1.23 (1.15-1.31, p = 0.000)** | **1.23 (1.04-1.46, p = 0.015)** | **1.52 (1.29-1.78, p = 0.000)** |
|  | Adjusted | **1.17 (1.08-1.26, p = 0.000)** | **1.23 (1.03-1.46, p = 0.026)** | **1.43 (1.20-1.70, p = 0.000)** |
|  | Adjusted - adding site | **1.09 (1.01-1.19, p = 0.027)** | **1.21 (1.01-1.44, p = 0.038)** | **1.32 (1.11-1.58, p = 0.002)** |
| **Chronic cough (wet)** | Crude | 1.12 (0.84-1.50, p = 0.443) | 1.05 (0.69-1.60, p = 0.811) | 1.18 (0.76-1.84, p = 0.464) |
|  | Adjusted | 1.11 (0.81-1.52, p = 0.515) | 1.05 (0.68-1.60, p = 0.838) | 1.16 (0.73-1.85, p = 0.533) |
|  | Adjusted - adding site | 1.03 (0.72-1.48, p = 0.858) | 1.04 (0.67-1.60, p = 0.868) | 1.07 (0.65-1.78, p = 0.788) |
| **Chronic cough (dry)** | Crude | 1.20 (0.93-1.56, p = 0.162) | 1.35 (0.90-2.03, p = 0.148) | **1.62 (1.07-2.47, p = 0.024)** |
|  | Adjusted | 1.10 (0.81-1.48, p = 0.546) | 1.36 (0.88-2.10, p = 0.165) | 1.49 (0.94-2.37, p = 0.091) |
|  | Adjusted - adding site | 1.04 (0.74-1.47, p = 0.814) | 1.36 (0.88-2.10, p = 0.172) | 1.41 (0.86-2.31, p = 0.169) |
| **Current wheeze** | Crude | **1.19 (1.12-1.27, p = 0.000)** | **1.20 (1.02-1.42, p = 0.029)** | **1.44 (1.23-1.69, p = 0.000)** |
|  | Adjusted | **1.17 (1.09-1.26, p = 0.000)** | 1.19 (0.99-1.43, p = 0.065) | **1.39 (1.16-1.66, p = 0.000)** |
|  | Adjusted - adding site | 1.05 (0.97-1.14, p = 0.208) | 1.16 (0.97-1.40, p = 0.109) | **1.22 (1.02-1.47, p = 0.030)** |
| **Chest tightness** | Crude | **1.17 (1.08-1.26, p = 0.000)** | 1.03 (0.86-1.23, p = 0.773) | **1.20 (1.01-1.43, p = 0.041)** |
|  | Adjusted | **1.12 (1.02-1.23, p = 0.014)** | 0.99 (0.81-1.20, p = 0.886) | 1.10 (0.91-1.34, p = 0.310) |
|  | Adjusted - adding site | 1.04 (0.95-1.14, p = 0.409) | 0.97 (0.80-1.18, p = 0.776) | 1.01 (0.83-1.23, p = 0.913) |
| **Shortness of breath (nocturnal)** | Crude | 1.13 (0.89-1.45, p = 0.313) | 1.34 (0.90-2.01, p = 0.154) | 1.52 (1.00-2.32, p = 0.051) |
|  | Adjusted | 1.05 (0.80-1.37, p = 0.733) | 1.34 (0.88-2.04, p = 0.172) | 1.40 (0.89-2.21, p = 0.142) |
|  | Adjusted - adding site | 0.97 (0.71-1.33, p = 0.868) | 1.34 (0.88-2.05, p = 0.176) | 1.31 (0.81-2.12, p = 0.277) |
| **Shortness of breath (resting)** | Crude | 1.22 (0.96-1.55, p = 0.107) | 0.95 (0.64-1.40, p = 0.787) | 1.15 (0.77-1.72, p = 0.481) |
|  | Adjusted | 1.11 (0.85-1.46, p = 0.433) | 0.91 (0.61-1.37, p = 0.656) | 1.02 (0.66-1.57, p = 0.945) |
|  | Adjusted - adding site | 0.99 (0.72-1.35, p = 0.934) | 0.89 (0.59-1.36, p = 0.595) | 0.88 (0.55-1.42, p = 0.602) |
| **Current nasal symptoms** | Crude | 1.03 (0.97-1.10, p = 0.383) | 0.96 (0.82-1.14, p = 0.664) | 0.99 (0.85-1.16, p = 0.920) |
|  | Adjusted | 1.06 (0.98-1.13, p = 0.130) | 0.95 (0.80-1.13, p = 0.600) | 1.01 (0.85-1.19, p = 0.921) |
|  | Adjusted - adding site | 1.03 (0.95-1.12, p = 0.424) | 0.95 (0.80-1.13, p = 0.572) | 0.98 (0.83-1.17, p = 0.843) |
| **Chronic phlegm** | Crude | 1.23 (0.97-1.55, p = 0.090) | 1.20 (0.82-1.75, p = 0.358) | 1.46 (0.99-2.17, p = 0.057) |
|  | Adjusted | 1.11 (0.85-1.46, p = 0.432) | 1.21 (0.80-1.83, p = 0.378) | 1.34 (0.86-2.09, p = 0.189) |
|  | Adjusted - adding site | 1.05 (0.77-1.44, p = 0.736) | 1.19 (0.78-1.81, p = 0.409) | 1.26 (0.78-2.02, p = 0.340) |

Table S3. Effects of 10µg/m^3^ increase in mean coalmine fire-related PM_2.5_ and COVID-19 on prevalence of respiratory symptoms and whether there is a moderating effect, 8.5-9 years after the coalmine fire survey; significant associations at p ≤ 0.05 in bold

| **Respiratory symptom** | **Model type** | **PM_2.5_ effect, no history of COVID-19** | **PM_2.5_ * COVID-19 effect** | **PM_2.5_ effect, history of COVID-19** |
| --- | --- | --- | --- | --- |
| **Chronic cough** | Crude | **1.34 (1.10-1.64, p = 0.004)** | 1.04 (0.77-1.41, p = 0.805) | **1.40 (1.11-1.76, p = 0.004)** |
|  | Adjusted | **1.34 (1.07-1.67, p = 0.010)** | 0.95 (0.69-1.31, p = 0.750) | 1.27 (0.99-1.62, p = 0.061) |
|  | Adjusted - adding site | 1.25 (0.98-1.60, p = 0.073) | 0.95 (0.69-1.32, p = 0.762) | 1.19 (0.91-1.55, p = 0.204) |
| **Chronic cough – wet** | Crude | 1.18 (0.95-1.47, p = 0.145) | 1.31 (0.94-1.83, p = 0.113) | **1.54 (1.20-1.98, p = 0.001)** |
|  | Adjusted | 1.20 (0.94-1.52, p = 0.141) | 1.22 (0.86-1.73, p = 0.274) | **1.46 (1.12-1.89, p = 0.005)** |
|  | Adjusted - adding site | 1.15 (0.88-1.50, p = 0.311) | 1.22 (0.86-1.75, p = 0.264) | **1.41 (1.06-1.85, p = 0.016)** |
| **Chronic cough – dry** | Crude | **1.27 (1.03-1.57, p = 0.027)** | 0.79 (0.55-1.13, p = 0.192) | 1.00 (0.76-1.33, p = 0.989) |
|  | Adjusted | 1.21 (0.95-1.54, p = 0.116) | 0.77 (0.53-1.12, p = 0.170) | 0.93 (0.69-1.26, p = 0.646) |
|  | Adjusted - adding site | 1.13 (0.87-1.47, p = 0.354) | 0.76 (0.51-1.12, p = 0.163) | 0.86 (0.61-1.20, p = 0.374) |
| **Current wheeze** | Crude | 1.17 (0.96-1.44, p = 0.127) | 1.16 (0.85-1.59, p = 0.343) | **1.37 (1.08-1.73, p = 0.010)** |
|  | Adjusted | 1.12 (0.90-1.41, p = 0.317) | 1.18 (0.84-1.64, p = 0.339) | **1.32 (1.02-1.71, p = 0.034)** |
|  | Adjusted - adding site | 1.15 (0.90-1.47, p = 0.265) | 1.14 (0.83-1.59, p = 0.419) | 1.32 (1.00-1.73, p = 0.051) |
| **Chest tightness** | Crude | 1.01 (0.82-1.24, p = 0.937) | 1.25 (0.92-1.69, p = 0.158) | **1.26 (1.01-1.56, p = 0.041)** |
|  | Adjusted | 0.95 (0.76-1.20, p = 0.684) | 1.32 (0.95-1.81, p = 0.094) | 1.25 (0.99-1.59, p = 0.060) |
|  | Adjusted - adding site | 0.90 (0.70-1.17, p = 0.448) | 1.31 (0.95-1.80, p = 0.101) | 1.18 (0.92-1.52, p = 0.191) |
| **Shortness of breath (nocturnal)** | Crude | 1.10 (0.88-1.37, p = 0.403) | 1.27 (0.92-1.74, p = 0.145) | **1.39 (1.11-1.75, p = 0.005)** |
|  | Adjusted | 1.02 (0.81-1.29, p = 0.854) | 1.31 (0.95-1.82, p = 0.102) | **1.34 (1.05-1.71, p = 0.018)** |
|  | Adjusted - adding site | 0.91 (0.70-1.18, p = 0.479) | 1.32 (0.94-1.86, p = 0.104) | 1.20 (0.93-1.57, p = 0.167) |
| **Shortness of breath (resting)** | Crude | **1.27 (1.04-1.55, p = 0.019)** | 0.82 (0.60-1.12, p = 0.208) | 1.04 (0.83-1.32, p = 0.726) |
|  | Adjusted | 1.23 (0.99-1.52, p = 0.066) | 0.81 (0.59-1.12, p = 0.205) | 1.00 (0.78-1.28, p = 0.977) |
|  | Adjusted - adding site | 1.18 (0.93-1.50, p = 0.164) | 0.80 (0.58-1.11, p = 0.180) | 0.95 (0.72-1.25, p = 0.704) |
| **Current nasal symptoms** | Crude | 0.90 (0.72-1.12, p = 0.348) | 1.06 (0.76-1.47, p = 0.733) | 0.95 (0.75-1.22, p = 0.702) |
|  | Adjusted | 0.89 (0.71-1.13, p = 0.342) | 1.07 (0.77-1.50, p = 0.679) | 0.96 (0.74-1.25, p = 0.762) |
|  | Adjusted - adding site | 0.83 (0.64-1.08, p = 0.159) | 1.07 (0.76-1.49, p = 0.703) | 0.89 (0.67-1.18, p = 0.411) |
| **Chronic phlegm** | Crude | 1.10 (0.89-1.37, p = 0.368) | 1.32 (0.95-1.83, p = 0.099) | **1.46 (1.15-1.85, p = 0.002)** |
|  | Adjusted | 1.10 (0.87-1.39, p = 0.444) | 1.23 (0.88-1.74, p = 0.227) | **1.35 (1.05-1.74, p = 0.019)** |
|  | Adjusted - adding site | 1.00 (0.77-1.30, p = 0.997) | 1.24 (0.87-1.75, p = 0.230) | 1.24 (0.94-1.62, p = 0.124) |

### 5 Sensitivity analyses

#### 5.1 Asthma-stratified analyses of changes in respiratory symptoms

Longitudinal analyses of respiratory symptoms suggested coalmine fire-related PM_2.5_ exposure increased the prevalence of current wheeze. Earlier Hazelwood Health Study analyses found limited evidence that Morwell residents had poorer asthma control, though there was no detectable association with PM_2.5_. The authors postulated that poorer asthma control in Morwell could nevertheless be explained by asthma being exacerbated by smoke exposure.^2^

Using self-reported asthma at the initial 2016/17 survey, we conducted stratified longitudinal analyses of PM_2.5_ exposure’s effect on respiratory symptoms. The results suggest effects were mostly isolated to non-asthmatics, including increased prevalence of chronic cough, current wheeze, chest tightness, and chronic phlegm. There was also a reduction in chest tightness among asthmatics, which could be due to increased use in asthma medications.

The asthmatic subsample had less statistical power due to smaller numbers (*n* = 999 at the initial 2016/17 survey and 176 at the 2022 follow-up, compared to *n* = 3,042 and *n* = 433 among non-asthmatics). While this could mean that effects were harder to detect within asthmatics, point estimates indicate the exacerbation of respiratory symptoms was isolated to non-asthmatics. As we note in the *Discussion* of the main document, this may be because asthmatics have access to inhaled medications like corticosteroids that could control respiratory symptoms brought on by the coalmine fire. For instance, majority of asthmatics (*n* = 601, 60.2%) reported taking some sort of medication, though we have no further details about kind or frequency. Medication access may also explain why chest tightness in decreased in asthmatics between the initial and follow-up surveys. These findings are illustrated in Figure S1.

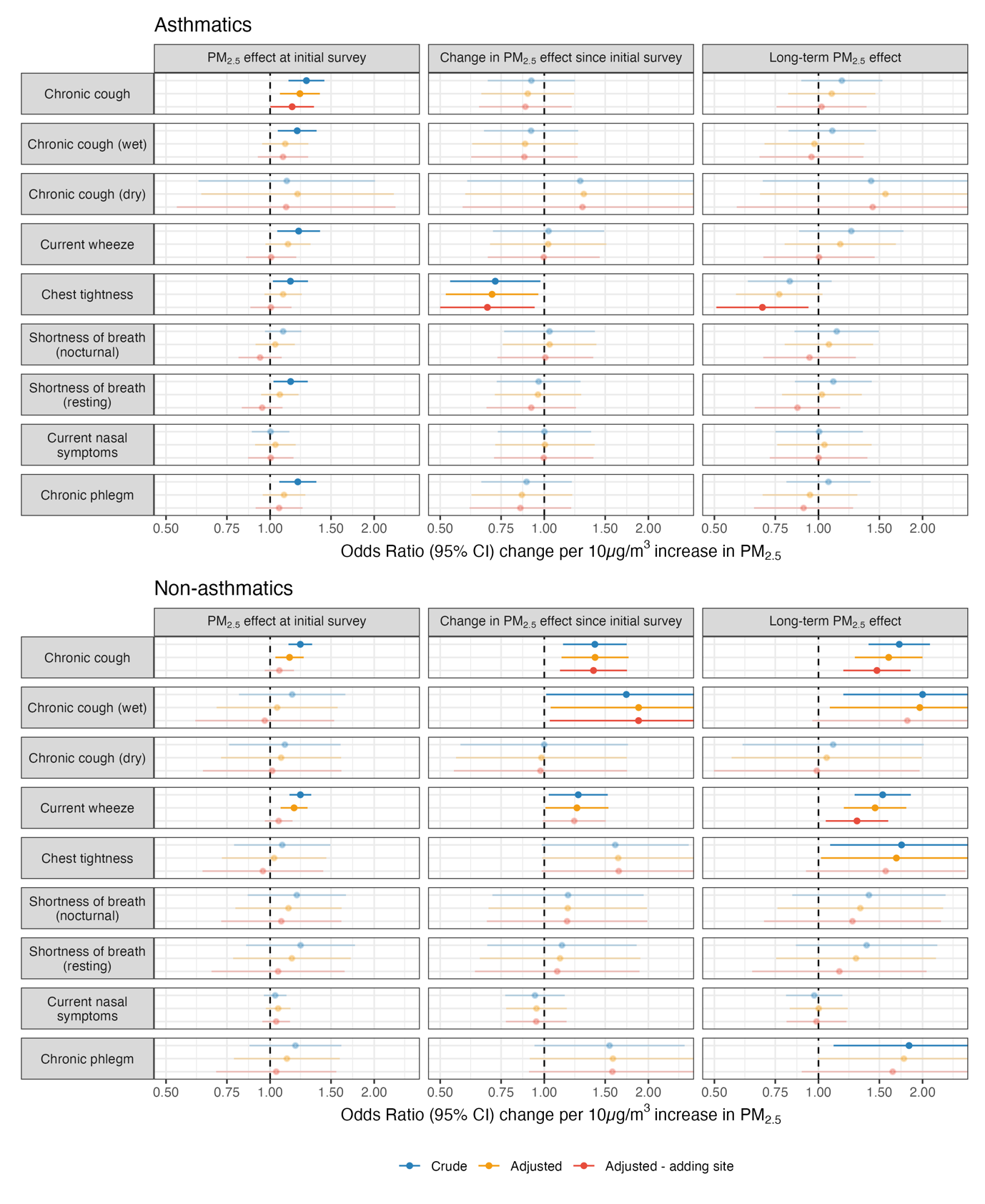

Figure S2. Analyses stratified by self-reported by asthma status on effects of 10µg/m^3^ increase in mean coalmine fire-related PM_2.5_ on odds of respiratory symptom at the initial 2016/17 survey (2-3 years post-fire), change between the two survey rounds (interaction term between fire-related PM_2.5_ and survey round), and the cumulative long-term effect at the 2022 follow-up (8.5-9 years post-fire, estimated using linear combination); faded points and intervals indicate non-significant effects

#### 5.2 COVID-19 * PM_2.5_ analyses using inverse probability weights

Invited cohort members who took part in the follow-up were more likely to be female (*p*=0.032), younger (*p*=0.002), in better health (*p* = 0.017), have asthma (*p*=0.014), and more likely to be a former smoker than current smoker (*p* < 0.001), but were less likely to have COPD (*p*=0.002) before the coalmine fire, or be more highly educated (*p* < 0.001) or from a higher socioeconomic area (*p* < 0.001). These analyses are summarised in Table S2. These factors were incorporated into inverse probability weighting to account for attrition bias.

Table S4. Attrition analyses comparing non-participants and participants in the follow-up survey; comparisons made with Fisher’s exact test for dichotomous and categorical variables and Wilcoxon rank-sum test for continuous variables

|  | **Non-participating (*n* = 1,846)** | **Participating (*n* = 612)** | ***p*-value** |
| --- | --- | --- | --- |
| **Daily mean fire-related PM_2.5_ exposure** | 8µg/m^3^ [IQR: 0, 14] | 8µg/m^3^ [IQR: 0, 14] | 0.338 |
| **Demographics** | | | |
| **Age at survey**  ***Missing*** | 60 [IQR: 47, 69]  *2* | 58 [IQR: 49, 65]  *1* | **0.003** |
| **Female** | 846 (46%) | 250 (41%) | **0.032** |
| **Residential SES***  ***Missing*** | 844 [IQR: 806, 930]  *27* | 869 [IQR: 814, 936]  *14* | **< 0.001** |
| **Educational attainment**  **Secondary to year 10**  **Secondary to year 11-12**  **Tertiary/trade qualification**  ***Missing*** | 584 (32%)  377 (21%)  868 (47%)  *17* | 127 (21%)  121 (20%)  361 (59%)  *3* | **< 0.001** |
| **Smoker**  **Non-smoker**  **Previous smoker**  **Current smoker**  ***Missing*** | 873 (47%)  623 (34%)  345 (19%)  *5* | 286 (47%)  249 (41%)  74 (12%)  *3* | **< 0.001** |
| **Cigarette pack-years**  ***Missing*** | 0 [IQR: 0, 15]  *42* | 0 [IQR: 0, 14]  *10* | 0.869 |
| **Health** | | | |
| **General health**  **Excellent**  **Very good**  **Good**  **Fair**  **Poor**  ***Missing*** | 189 (10%)  516 (28%)  630 (34%)  352 (19%)  145 (7.9%)  *14* | 67 (11%)  199 (33%)  220 (36%)  95 (16%)  31 (5.1%)  *0* | **0.017** |
| **Pre-2014 asthma**  ***Missing*** | 393 (21%)  *7* | 160 (26%)  *1* | **0.014** |
| **Pre-2014 COPD**  ***Missing*** | 93 (5.0%)  *3* | 13 (2.1%)  *2* | **0.002** |

*From the Index of Relative Socio-economic Advantage and Disadvantage (IRSAD)^1^

When inverse probability weights were added to analyses, the precision of estimates was reduced, as indicated by wider confidence intervals, and the effect of PM_2.5_ on shortness of breath (resting) among those with no history of COVID-19 attenuated substantially. Otherwise, the direction and magnitude of effects remained similar, if less frequently statistically significant. These results are summarised in Figure S3 and Table S5.

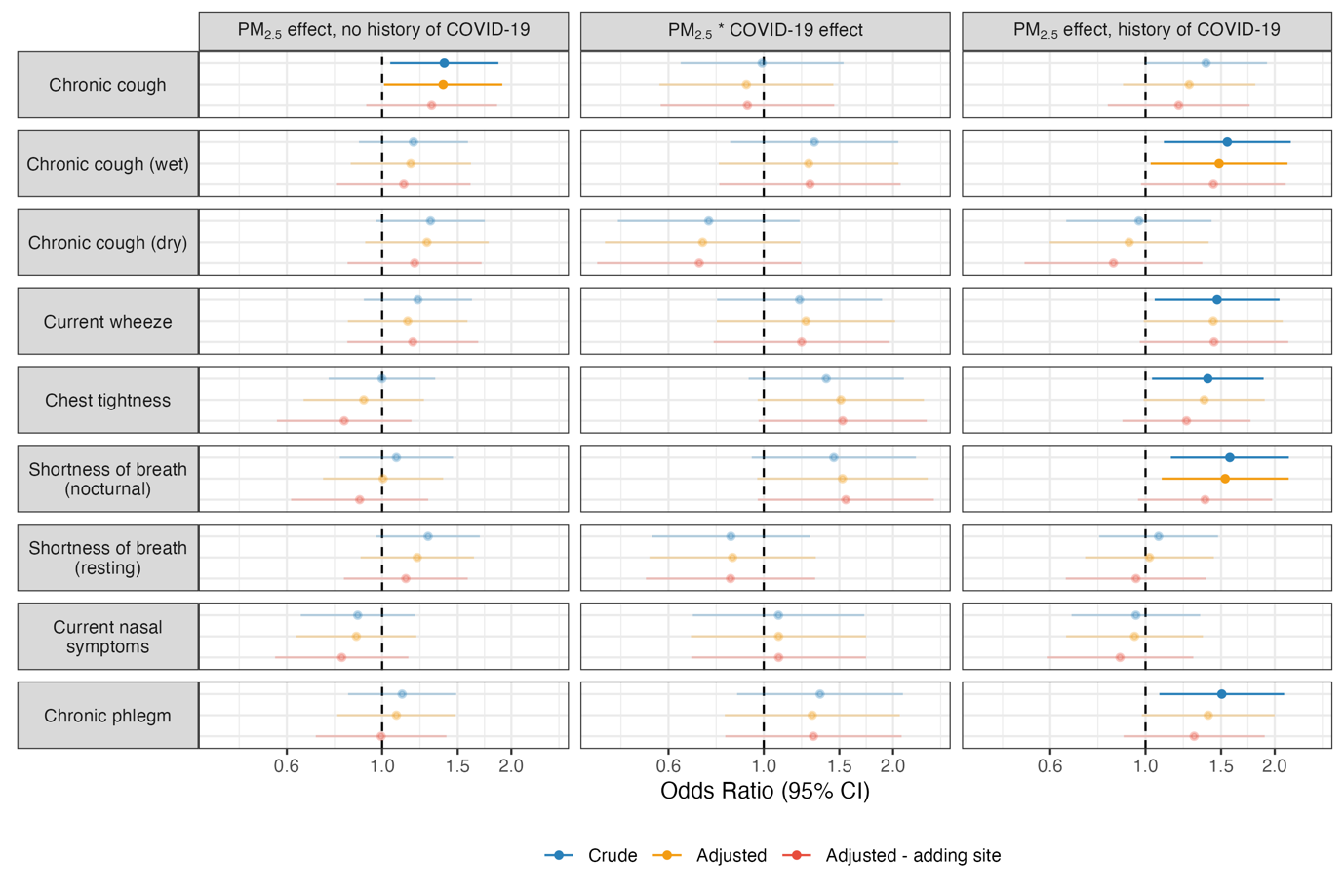
Figure S3. Effects of 10µg/m^3^ increases in coalmine fire-related PM_2.5_ and COVID-19 on prevalence of respiratory symptoms and whether there is a moderating effect, 8.5-9 years after the fire; faded points and intervals indicate non-significant effects – weighted for attrition bias using inverse probability weighting

Table S5. Effects of 10µg/m^3^ increase in mean coalmine fire-related PM_2.5_ and COVID-19 on prevalence of respiratory symptoms and whether there is a moderating effect, 8.5-9 years after the coalmine fire survey; significant associations at p ≤ 0.05 in bold – weighted for attrition bias using inverse probability weighting

| **Respiratory symptom** | **Model type** | **PM_2.5_ effect, no history of COVID-19** | **PM_2.5_ * COVID-19 effect** | **PM_2.5_ effect, history of COVID-19** |
| --- | --- | --- | --- | --- |
| **Chronic cough** | Crude | **1.40 (1.04-1.87, p = 0.024)** | 0.99 (0.64-1.53, p = 0.968) | 1.38 (1.00-1.92, p = 0.052) |
|  | Adjusted | **1.39 (1.01-1.90, p = 0.043)** | 0.91 (0.57-1.45, p = 0.695) | 1.26 (0.89-1.80, p = 0.196) |
|  | Adjusted - adding site | 1.30 (0.92-1.85, p = 0.138) | 0.92 (0.58-1.46, p = 0.712) | 1.19 (0.82-1.75, p = 0.359) |
| **Chronic cough – wet** | Crude | 1.18 (0.88-1.58, p = 0.259) | 1.31 (0.84-2.06, p = 0.239) | **1.55 (1.10-2.18, p = 0.011)** |
|  | Adjusted | 1.17 (0.85-1.61, p = 0.349) | 1.27 (0.79-2.06, p = 0.328) | **1.48 (1.03-2.14, p = 0.035)** |
|  | Adjusted - adding site | 1.12 (0.79-1.61, p = 0.525) | 1.28 (0.79-2.08, p = 0.318) | 1.44 (0.98-2.12, p = 0.066) |
| **Chronic cough – dry** | Crude | 1.30 (0.97-1.74, p = 0.081) | 0.74 (0.46-1.21, p = 0.235) | 0.97 (0.65-1.43, p = 0.858) |
|  | Adjusted | 1.27 (0.91-1.77, p = 0.154) | 0.72 (0.43-1.22, p = 0.219) | 0.92 (0.60-1.40, p = 0.687) |
|  | Adjusted - adding site | 1.19 (0.83-1.71, p = 0.342) | 0.71 (0.41-1.22, p = 0.215) | 0.84 (0.52-1.36, p = 0.480) |
| **Current wheeze** | Crude | 1.21 (0.91-1.62, p = 0.194) | 1.21 (0.78-1.89, p = 0.395) | **1.47 (1.05-2.05, p = 0.025)** |
|  | Adjusted | 1.15 (0.83-1.58, p = 0.402) | 1.25 (0.78-2.02, p = 0.354) | 1.44 (0.99-2.09, p = 0.056) |
|  | Adjusted - adding site | 1.18 (0.83-1.67, p = 0.358) | 1.22 (0.76-1.96, p = 0.399) | 1.44 (0.97-2.15, p = 0.071) |
| **Chest tightness** | Crude | 1.00 (0.75-1.33, p = 0.996) | 1.40 (0.92-2.12, p = 0.115) | **1.40 (1.04-1.88, p = 0.029)** |
|  | Adjusted | 0.91 (0.66-1.25, p = 0.550) | 1.51 (0.97-2.36, p = 0.069) | 1.37 (0.99-1.90, p = 0.057) |
|  | Adjusted - adding site | 0.82 (0.57-1.17, p = 0.270) | 1.53 (0.97-2.40, p = 0.066) | 1.25 (0.88-1.76, p = 0.210) |
| **Shortness of breath (nocturnal)** | Crude | 1.08 (0.80-1.46, p = 0.621) | 1.46 (0.94-2.26, p = 0.094) | **1.57 (1.15-2.16, p = 0.005)** |
|  | Adjusted | 1.01 (0.73-1.39, p = 0.974) | 1.53 (0.97-2.41, p = 0.070) | **1.53 (1.09-2.16, p = 0.014)** |
|  | Adjusted - adding site | 0.89 (0.61-1.28, p = 0.521) | 1.55 (0.97-2.49, p = 0.068) | 1.38 (0.96-1.97, p = 0.082) |
| **Shortness of breath (resting)** | Crude | 1.28 (0.97-1.69, p = 0.082) | 0.84 (0.55-1.28, p = 0.414) | 1.07 (0.78-1.48, p = 0.666) |
|  | Adjusted | 1.21 (0.89-1.64, p = 0.224) | 0.85 (0.54-1.32, p = 0.462) | 1.02 (0.72-1.44, p = 0.903) |
|  | Adjusted - adding site | 1.14 (0.81-1.58, p = 0.453) | 0.84 (0.53-1.32, p = 0.441) | 0.95 (0.65-1.38, p = 0.790) |
| **Current nasal symptoms** | Crude | 0.88 (0.65-1.19, p = 0.401) | 1.08 (0.68-1.71, p = 0.736) | 0.95 (0.67-1.34, p = 0.769) |
|  | Adjusted | 0.87 (0.63-1.20, p = 0.400) | 1.08 (0.68-1.73, p = 0.741) | 0.94 (0.65-1.36, p = 0.753) |
|  | Adjusted - adding site | 0.81 (0.56-1.15, p = 0.236) | 1.08 (0.68-1.73, p = 0.738) | 0.87 (0.59-1.29, p = 0.496) |
| **Chronic phlegm** | Crude | 1.11 (0.83-1.49, p = 0.467) | 1.35 (0.87-2.11, p = 0.184) | **1.51 (1.08-2.10, p = 0.017)** |
|  | Adjusted | 1.08 (0.79-1.48, p = 0.635) | 1.30 (0.81-2.07, p = 0.277) | 1.40 (0.98-2.00, p = 0.064) |
|  | Adjusted - adding site | 0.99 (0.70-1.41, p = 0.976) | 1.31 (0.81-2.09, p = 0.270) | 1.30 (0.89-1.90, p = 0.177) |

### 6 Respiratory symptom questions

Respiratory symptom items were derived the *European Community Respiratory Health Survey III – Screening Questionnaire*^3^ and are reproduced here.

- **Chronic cough:** Do you cough on most days for as much as three months a year?
- **Current wheeze:** Have you had wheezing or whistling in your chest at any time in the last 12 months?
- **Chest tightness:** Have you woken up with a feeling of tightness in your chest at any time in the last 12 months?
- **Shortness of breath (nocturnal):** Have you been woken by an attack of shortness of breath at any time in the last 12 months?
- **Shortness of breath (resting):** Have you had an attack of shortness of breath that came on during the day when you were at rest at any time in the last 12 months?
- **Current nasal symptoms:** Do you have any nasal allergies including hay fever?
- **Chronic phlegm:** Do you bring up phlegm from your chest on most days for as much as three months a year?

Wet chronic cough was determined by presence of both chronic cough and chronic phlegm, whereas dry chronic cough was determined by presence of chronic cough and *absence* of chronic phlegm.

### 7 Deviations from pre-registered analysis plan

This study was pre-registered on the Open Science Framework on 19 July 2022, the month before the follow-up survey commenced, and updated during the survey with a more specific analysis plan on 22 November 2022.^4^

We initially planned to include terms for PM_2.5_ exposure from the 2019/2020 Black Summer bushfire, although the necessary modelled data were not available as of writing. Additionally, we found there was likely little difference in exposure between Morwell and Sale.^5,6^ Research questions were clarified to reflect this change.

Initially, we intended to test two approaches for tobacco use (smoker status versus pack-years to capture) and select one based on model fit, but decided to use both. The square root transformation of pack years was not pre-specified, but applied after observing the extreme right-skewed in the distributions in a prior analysis.^6^ While age was pre-specified as age group, we opted to use the natural spline to better account for non-linear effects while also retaining the rich information of a continuous rather than recoding into categorical variable. Body Mass Index was excluded as a confounder because it only available for those who participated in the follow-up survey.

### 8 Additional resources

#### 8.1 Statistical packages for cleaning and analysing study data

To give credit to authors of the statistical packages used in this paper, we have listed theme here. All analyses were conducted in *R*^7^ in the *RStudio* IDE.^8^ Specific packages include: *broom.mixed*^9^ to produce tidy results tables; *glmmTMB*^10^ for mixed-effect logistic regression modelling; *gtsummary*^11^ for descriptive tables and univariate comparisons; *haven*^12^ to import data from different statistical package formats; *janitor*^13^ to clean column names; *mice*^14^ for multiple imputation; *multcomp*^15^ to calculate the linear combination of effects and confidence intervals from interaction terms; *naniar*^16^ to analyse missingness, patchwork^17^ to combine plots; *readxl*^18^ to import excel file data; and *tidyverse*^19^ to use the suite of tidyverse tools for importation, manipulation, and visualisation tools. All cleaning and analytical code have been archived on a public repository.^20^

#### 8.2 Mapping data and software

To create the map of coalmine fire-related PM_2.5_ distribution in Figure 1, we used geospatial data of national, state/territory, and Statistical Area Boundaries from the Australian Bureau of Statistics,^21^ coalmine boundaries from the Victorian Department of Primary Industries^22^ and Department of Energy, Environment and Climate Action’s DataShare platform,^23^ and modelled PM_2.5_ distribution estimates.^24^ The map was created using the following R packages: *rmapshaper*^25^ to simplify map boundaries, *sf*^26,27^ to read and write shapefiles, and *tmap*^28^ to draw and save maps.

22	Department of Primary Industries. Victorian Coal: A 2006 Inventory of Resources. Morwell, Victoria: GHD Pty Ltd, 2007.

23	Department of Energy, Environment and Climate Action. Victorian Coal Fields. 2007; published online Aug 31. https://datashare.maps.vic.gov.au.

24	Luhar AK, Emmerson KM, Reisen F, Williamson GJ, Cope ME. Modelling smoke distribution in the vicinity of a large and prolonged fire from an open-cut coal mine. *Atmos Environ* 2020; **229**: 117471.

25	Teucher A, Russell K. rmapshaper: Client for ‘mapshaper’ for ‘Geospatial’ Operations. 2023. https://CRAN.R-project.org/package=rmapshaper.

26	Pebesma E. Simple Features for R: Standardized Support for Spatial Vector Data. *R J* 2018; **10**: 439–46.

27	Pebesma E, Bivand R. Spatial Data Science: With Applications in R. New York: Chapman and Hall/CRC, 2023 https://doi.org/10.1201/9780429459016.

28	Tennekes M. tmap: Thematic Maps in R. *J Stat Software2* 2018; **84**: 1–39.
